# Newborn Differential DNA Methylation and Subcortical Brain Volumes as Early Signs of Severe Neurodevelopmental Delay in a South African Birth Cohort Study

**DOI:** 10.1101/2020.03.13.20035501

**Authors:** Anke Hüls, Catherine J Wedderburn, Nynke A Groenewold, Nicole Gladish, Meaghan Jones, Nastassja Koen, Julia L MacIsaac, David TS Lin, Katia E Ramadori, Michael P Epstein, Kirsten A. Donald, Michael S Kobor, Heather J Zar, Dan J Stein

**Affiliations:** Department of Epidemiology and Gangarosa Department of Environmental Health, Rollins School of Public Health, Emory University, Atlanta, Georgia, USA; Department of Paediatrics and Child Health, Red Cross War Memorial Children’s Hospital, University of Cape Town, SA; Department of Clinical Research, London School of Hygiene & Tropical Medicine, London, UK; Neuroscience Institute, University of Cape Town, Cape Town, South Africa; Department of Psychiatry and Mental Health, University of Cape Town, Cape Town, South Africa; Department of Medical Genetics, University of British Columbia, Vancouver, BC, Canada; BC Children’s Hospital Research Institute, Vancouver, BC, Canada; Centre for Molecular Medicine and Therapeutics, Vancouver, BC, Canada; Department of Biochemistry and Medical Genetics, University of Manitoba, and Children’s Hospital Research, Institute of Manitoba, Winnipeg, Canada; South African Medical Research Council (SAMRC) Unit on Risk and Resilience in Mental Disorders, University of Cape Town, Cape Town, South Africa; Department of Human Genetics, School of Medicine, Emory University, Atlanta, Georgia, USA; South African Medical Research Council (SAMRC) Unit on Child and Adolescent Health, University of Cape Town, Cape Town, South Africa

**Author notes:** **Corresponding author:** Anke Huels, PhD, Rollins School of Public Health, Emory University, 1518 Clifton Road, Atlanta, GA 30322. **Competing financial interests declaration:** All authors declare they have no actual or potential competing financial interest.

## Abstract

**Objective:** The first two years of life are a critical period of rapid brain development. Since early neurodevelopment is influenced by prenatal risk factors and genetics, neonatal biomarkers can potentially provide the opportunity to detect early signs of neurodevelopmental delay. We analyzed associations between DNA methylation (DNAm) levels from cord blood, neonatal magnetic resonance imaging (MRI) neuroimaging data, and neurodevelopment at two years of age.

**Methods:** Neurodevelopment was assessed in 161 children from the South African Drakenstein Child Health Study at two years of age using the Bayley Scales of Infant and Toddler Development III. We performed an epigenome-wide association study of neurodevelopmental delay using DNAm levels from cord blood. A mediation analysis was conducted in 51 children to analyze if associations between differential DNAm and neurodevelopmental delay were mediated by altered neonatal brain volumes.

**Results:** We found epigenome-wide significant associations between differential DNAm at the *SPTBN4* locus (cg26971411, p-value=3.10×10^−08^), an intergenic region on chromosome 11 (cg00490349, p-value=2.41×10^−08^) and a differentially methylated region on chromosome 1 (FDR p-value for the region=9.06×10^−05^) and severe neurodevelopmental delay. While these associations were not mediated by neonatal brain volume, neonatal caudate volumes were independently associated with neurodevelopmental delay, particularly in language (p=0.0443) and motor (p=0.0082) domains.

**Conclusion:** Differential DNAm levels from cord blood and increased neonatal caudate volumes were independently associated with severe neurodevelopmental delay at two years of age. These findings suggest that neurobiological signals for severe developmental delay may be detectable in very early life with implications for identification and intervention design.

## Introduction

The first two years of life are a critical period of rapid growth and brain development. Child cognitive development is influenced by genetic and environmental factors which interact to determine how the brain develops and functions^1^. Multiple factors have been shown to affect neurodevelopment; these include poverty, maternal education, maternal physical and psychological health, substance use, and nutrition ^2–6^. As a result, low- and middle-income countries in general, and in particular sub-Saharan Africa, have a very high proportion of young children at risk of developmental delay^3^. New research in early human development shows that epigenetic, immunological, physiological, and psychological adaptations to the environment occur from conception, and that these adaptations affect development throughout the life course^6^. This highlights the importance of early identification of developmental risk and timely implementation of targeted interventions^4^. However, screening for neurodevelopmental delay is inconsistent across different environments, and in low-resource contexts where children are at highest risk, often does not occur in a rigorous manner^7^. Therefore, exploring neonatal neurobiological signals that can potentially be used to detect early signs of severe neurodevelopmental delay could have implications for identification and intervention design.

As such, DNA methylation (DNAm) levels from cord blood could potentially be used as neurobiological signals to understand future disease risk. DNAm can be altered by psychological, environmental and genetic factors, and is one of the most studied modifications of the genome. Multiple studies have demonstrated that epigenetic modifications are associated with the risk of cognitive disability or impairment^8^. However, the role of epigenetic variation on early cognitive development of infants remains largely unknown. Neuropsychiatric outcomes that are known to be associated with altered DNAm in newborns include attention-deficit/hyperactivity disorder (ADHD)^9^ and amygdala:hippocampal volume ratio, which is a marker for anxiety and aggression in later years^10^. Some evidence that altered methylation levels in neonates can also be used as a biomarker for neurodevelopmental delay comes from a study of 238 Mexican-American children^11^ which found non-significant associations between two methylation sites and working memory and processing speed.

Over the last decades, structural magnetic resonance imaging (MRI) studies in children have increasingly been used to ascertain relations between alteration in regional brain volumes and various functional measures of behavioral development^12,13^. Changes in brain volume have been associated with developmental disorders such as autism spectrum disorders^14,15^, ADHD^16^, and specific language impairment^17^. However, most of these studies have been conducted in school-aged children and information regarding structural and functional brain development in the earliest years of life remains limited and is mainly focused on premature infants^18,19^. More recent studies have used MRI brain scans from neonates and infants at 6 or 7 months of age to predict cognitive development, suggesting involvement of the maturation of subcortical brain regions^13,20^. Given their association with prenatal environmental, psychological risk factors and genetics^6^, we hypothesize that early changes in brain volume can potentially mediate the association between DNAm and neurodevelopmental delay.

In this study we will investigate associations between DNAm levels from cord blood, subcortical volumes from neonatal MRI imaging data and neurodevelopmental delay at two years of age in the South African Drakenstein Child Health Study (DCHS).

## Methods

### Study design and study population

The DCHS, a population-based birth cohort, has been described previously^21^. Mothers were enrolled prenatally in their second trimester and followed through pregnancy at two primary care clinics serving two distinct populations (predominantly black African ancestry or predominantly mixed ancestry). Mother-child pairs were followed from birth and infants enrolled in the DCHS were followed until at least five years of age^21^. All births occurred at a single, central facility, Paarl Hospital. The sample included in the present study were 161 children who had all measurements available, including cognitive Bayley Scales of Infant and Toddler Development, third edition (BSID III) scores at 2 years of age, methylation data from cord blood, genotyping data and all relevant covariates. A sub-set of 51 children also had neonatal MRI data available.

Ethical approval for human subjects’ research was obtained from the Human Research Ethics Committee of the Faculty of Health Sciences of University of Cape Town (HREC UCT REF 401/2009; HREC UCT REF 525/2012). Written informed consent was signed by the mothers on behalf of herself and her infant for participation in this study.

### DNA methylation

DNA was isolated from cord blood samples that were collected at time of delivery^22^. DNA methylation was assessed with the Illumina Infinium HumanMethylation450 BeadChips (n=156) and the MethylationEPIC BeadChips (n=160). Pre-processing and statistics were done using R 3.5.1^23^. Raw iDat files were imported to RStudio where intensity values were converted into beta values. The 450K and EPIC datasets were then combined using the minfi package^24^ resulting in 316 samples and 453,093 probes. Background subtraction, color correction and normalization were performed using the preprocessFunnorm function^25^. After sample and probe filtering, 273 samples and 409,033 probes remained for downstream analyses (see supplementary methods for details). Batch effects were removed using ComBat from the R package sva^26^. Cord blood cell type composition was predicted using the most recent cord blood reference data set^27^ and the IDOL algorithm and probe selection^28^.

### Genotype data

Genome-wide genotyping was performed in 305 newborns using the Illumina Infinium PsychArray (n=150) and the Illumina Infinium Global Screening Array, GSA (n=155), of which 271 samples passed quality control (QC). After QC SNPs were imputed on the 1000 Genomes reference panel (Phase III) using the Michigan Imputation Server^29^. Imputed genotypes reaching an R^2^≥ 0.3 in both arrays were used in analyses. Principal components were calculated using PLINK v1.90b4 64-bit.

### Assessment of neurodevelopmental delay

At 24 months, children were assessed using the BSID-III^30^. The BSID-III is a gold standard assessment of child development used internationally, and validated in South Africa^31^. The assessment generates scores for cognitive, language and motor development. Trained assessors administered the BSID-III using direct observation to score children on their cognitive, language and motor development^32^. BSID specialized software was used to produce normed and age-adjusted scores calculated using data from a US-reference group. Composite scores for cognitive, motor and language scales were scaled to have a mean of 100 and standard deviation of 15. The scores were categorized into severe delay if they were two standard deviations from the BSID-III reference mean, and mild delay if they were one standard deviation from the mean^30^.

### MRI imaging data from neonates

Neonatal imaging was performed on a subgroup of newborns from the DCHS at the Cape Universities Brain Imaging Centre, Tygerberg Hospital as has been described^2^. Newborns were excluded if they were found to have medical comorbidities or had neonatal intensive care admission at birth, were premature (<36 weeks’ gestation), had an APGAR core of <7 at 5 minutes, or their mothers used illicit drugs in pregnancy. Structural T2-weighted images were acquired during natural sleep on a Siemens Magnetom 3T Allegra MRI scanner (Erlangen, Germany) using a head coil with a wet clay inlay^33^. The acquisition parameters were: TR = 3500ms; TE = 354ms; FOV = 160×160mm; voxel dimensions = 1.3×1.3×1.0mm; 128 slices. Statistical parametric mapping software SPM8 (www.fil.ion.ucl.ac.uk/spm/software/spm8) was used to process the T2-images, using a well-established neonatal brain template^34^ for image normalization to standard space and segmentation of images into three tissue types using probabilistic maps as priors. Image alignment to the template and grey matter segmentations were visually inspected for accuracy. Volumetric data was extracted for the following subcortical regions: caudate, pallidum, putamen, thalamus, amygdala and hippocampus. The mean volume of the left and right hemispheres was used in all analyses. In addition, total grey matter, total white matter, and total cerebrospinal fluid (CSF) volumes were extracted and summed to obtain a measure of intracranial volume. More details regarding image acquisition and processing can be found elsewhere^35^. A subset of 51 of the 161 children had neonatal imaging data available for inclusion in this analysis.

### Statistical analysis

To identify DNA methylation patterns in cord blood that are associated with severe delay in neurodevelopment at two years of age, we conducted an epigenome-wide association study (EWAS) on single CpG sites, as well as an analysis of differentially methylated regions (DMRs). For the EWAS, we ran a multivariate robust linear regression model with empirical Bayes from the R package limma (version 3.40.6)^36^ using severe neurodevelopmental delay at the age of two years as the independent variable and methylation at each CpG site as a dependent variable, adjusting for sex, preterm birth, maternal smoking, household income, the first five genetic PCs to control for population stratification (see Figure S1 for details) and the first three PCs from the estimated cell type proportions (after centered log-ratio transformation), which explained >90% of the cell type heterogeneity^37^. The effect estimates from the adjusted models (**Δ** beta) refer to the difference in mean DNAm beta values between groups (with and without neurodevelopmental delay). We applied a Bonferroni threshold to correct for multiple testing based on the number of tested CpG sites (threshold: 0.05/403933=1.24 × 10^−07^). Fine-mapping of our epigenome-wide associations was done with coMET^38^, which is a visualization tool of EWAS results with functional genomic annotations and estimation of co-methylation patterns. We conducted the following sensitivity analyses: 1) We validated our findings by using mild delay in neurodevelopment and the continuous Bayley scores as independent variables, 2) We confirmed our associations using linear regression with p-values obtained from normal theory (lm() function in R), as well as from a permutation test. To identify plausible pathways associated with severe delay in neurodevelopment, we performed an over-representation analysis based on the CpGs with p-values < 0.001 for the association with severe delay in neurodevelopment. We used the R Bioconductor package missMethyl (version 1.18.0 gometh function), which performs one-sided hypergeometric tests, taking into account and correcting for any bias derived from the use of differing numbers of probes per gene interrogated by the array^39^. Differentially methylated regions (DMRs) in severe neurodevelopmental delay were identified using DMRcate, that identifies DMRs from tunable kernel smoothing processes of association signals^40^. Input files were our single-CpG EWAS results on severe neurodevelopmental delay including regression coefficients, standard deviations and uncorrected p-values. DMRs were defined based on the following criteria: a) a DMR should contain more than one probe; b) regional information can be combined from probes within 1,000 bp; c) the region showed FDR corrected p-value < 0.05.

Finally, we analyzed if associations between differential methylation and neurodevelopmental delay were mediated by neonatal brain volume as follows: (1) We analyzed associations between CpG sites, that were associated with neurodevelopmental delay, and MRI imaging data from neonates (total grey matter, total white matter and subcortical brain volumes) using linear regression models adjusted for the same covariates as the EWAS analyses, plus age at MRI scan, child sex and intracranial volume. (2) We analyzed associations between MRI imaging data and severe delay in neurodevelopment using linear regression models adjusted for age at MRI scan, child sex and intracranial volume. These associations were validated in two sensitivity analyses: First, associations were additionally adjusted for preterm birth, maternal smoking, household income and the first five genetic PCs to control for population stratification and second, the findings were validated by using mild neurodevelopmental delay and the continuous Bayley scores as independent variables. Lastly, if the associations in (1) and (2) were significant, a formal mediation analysis was conducted using the R package “mediation”, an approach that relies on the quasi-Bayesian Monte Carlo method based on normal approximation^41^.

## Results

### Description of Study Participants

The study sample included 161 children with DNA methylation and genotype data, BSID-III scores at two years and information on all relevant covariates (Table 1). Of these, neonatal MRI imaging data were available in 51 children and the average age at MRI scan was 3.1 weeks (sd = 0.9). Almost half of our study sample was female, half of them were of African ancestry and the other half were of mixed ancestry, and a quarter of the population was exposed to maternal smoking during pregnancy. Cognitive development at two years of age was severely delayed in 4 to 12% of the study sample, depending on the tested domain. The most prevalent delay was in language (8% in the whole study sample and 12% in the subsample with imaging data had severe language delay), followed by severe cognitive delay (7-8%) and severe delay in motor function (4%).

**Table 1.**
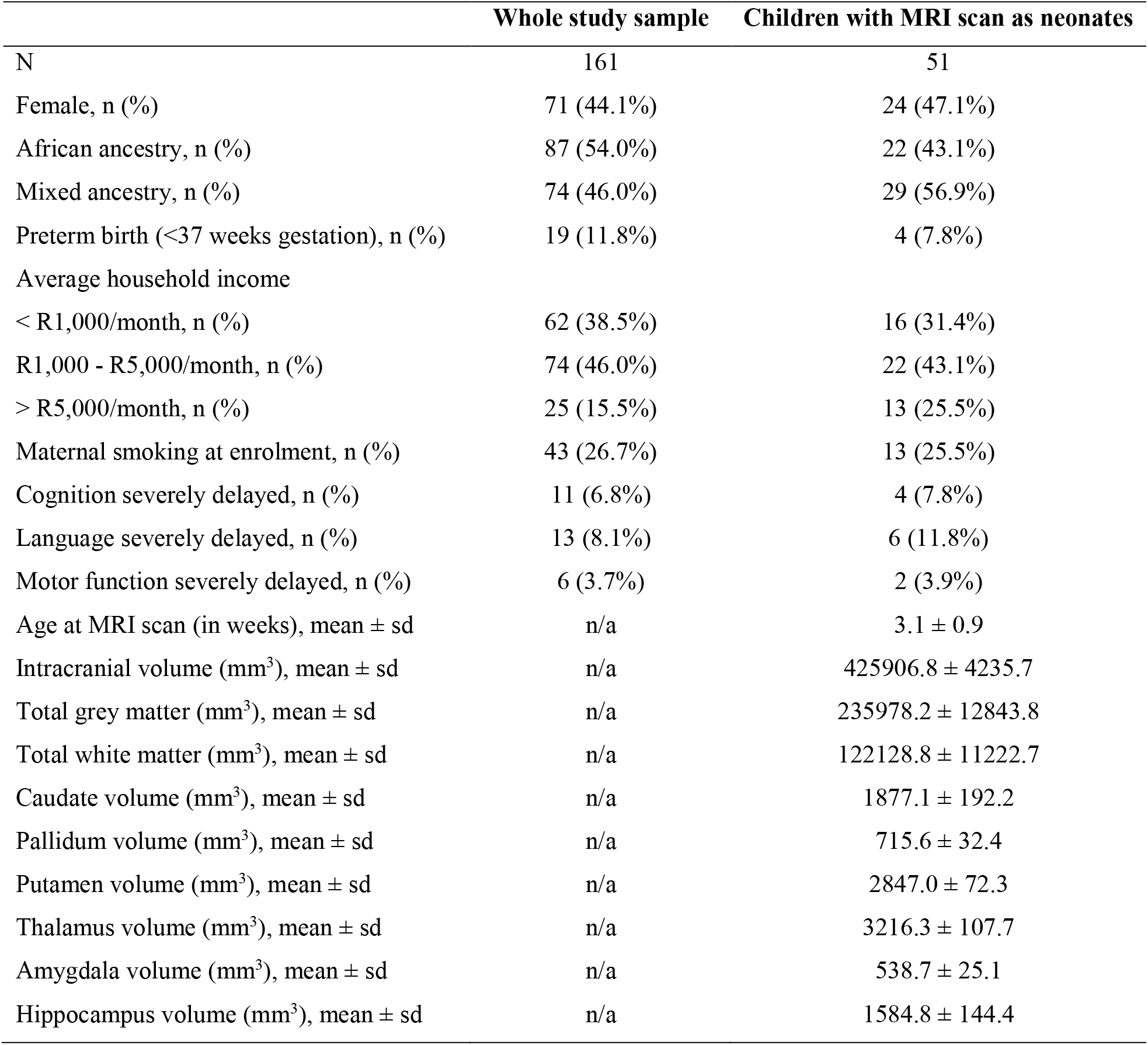
Study characteristics of the whole study population as well as the subsample with MRI imaging data.

|  | Whole study sample | Children with MRI scan as neonates |
| --- | --- | --- |
| N | 161 | 51 |
| Female, n (%) | 71 (44.1%) | 24 (47.1%) |
| African ancestry, n (%) | 87 (54.0%) | 22 (43.1%) |
| Mixed ancestry, n (%) | 74 (46.0%) | 29 (56.9%) |
| Preterm birth (<37 weeks gestation), n (%) | 19 (11.8%) | 4 (7.8%) |
| Average household income |  |  |
| < R1,000/month, n (%) | 62 (38.5%) | 16 (31.4%) |
| R1,000 - R5,000/month, n (%) | 74 (46.0%) | 22 (43.1%) |
| > R5,000/month, n (%) | 25 (15.5%) | 13 (25.5%) |
| Maternal smoking at enrolment, n (%) | 43 (26.7%) | 13 (25.5%) |
| Cognition severely delayed, n (%) | 11 (6.8%) | 4 (7.8%) |
| Language severely delayed, n (%) | 13 (8.1%) | 6 (11.8%) |
| Motor function severely delayed, n (%) | 6 (3.7%) | 2 (3.9%) |
| Age at MRI scan (in weeks), mean ± sd | n/a | 3.1 ± 0.9 |
| Intracranial volume (mm<sup>3</sup>), mean ± sd | n/a | 425906.8 ± 4235.7 |
| Total grey matter (mm<sup>3</sup>), mean ± sd | n/a | 235978.2 ± 12843.8 |
| Total white matter (mm<sup>3</sup>), mean ± sd | n/a | 122128.8 ± 11222.7 |
| Caudate volume (mm<sup>3</sup>), mean ± sd | n/a | 1877.1 ± 192.2 |
| Pallidum volume (mm<sup>3</sup>), mean ± sd | n/a | 715.6 ± 32.4 |
| Putamen volume (mm<sup>3</sup>), mean ± sd | n/a | 2847.0 ± 72.3 |
| Thalamus volume (mm<sup>3</sup>), mean ± sd | n/a | 3216.3 ± 107.7 |
| Amygdala volume (mm<sup>3</sup>), mean ± sd | n/a | 538.7 ± 25.1 |
| Hippocampus volume (mm<sup>3</sup>), mean ± sd | n/a | 1584.8 ± 144.4 |

### Differential Methylation as an Early Sign of Neurodevelopmental Delay

Differentially methylated CpG sites in the *SPTBN4* locus (cg26971411) and in an intergenic region on chromosome 11 (cg00490349) were associated with severe neurodevelopmental delay at the epigenome-wide significance level (Bonferroni-adjustment) after adjusting for sex, preterm birth, maternal smoking, household income, the first five genetic PCs and cell type proportions (Table 2, Figure 1, Figures S2 and S3). Differential methylation in cg26971411 was significantly associated with severe delay in motor function (**Δ** beta = −0.024, p-value = 3.10 × 10^−08^) and nominally significant for cognitive development (**Δ** beta = −0.013, p-value = 3.93 × 10^−05^) and language development (**Δ** beta = −0.014, p-value = 2.57 × 10^−06^). Differential methylation in cg00490349 was significantly associated with severe delay in language development (**Δ** beta = −0.036, p-value = 2.41 × 10^−08^) and nominally significant for cognitive development (**Δ** beta = −0.031, p-value = 1.94 × 10^−05^) and motor function (**Δ** beta = −0.050, p-value = 2.73 × 10^−07^). The significant associations were confirmed in a sensitivity analysis using linear regression models, as well as permutation tests (Table S1). Associations with mild neurodevelopmental delay were weaker, but still nominally significant, for differential methylation in cg26971411 and cg00490349 (p-values < 0.05, Table S2). Associations with the continuous Bayley scores were only nominally significant for cognitive function, but not for language development or motor function (Table S2).

**Table 2.** Significant single CpG sites and differentially methylated regions (DMRs) in relation to severe neurodevelopmental delay.

| A. Single CpGs |  |  |  | Cognition |  | Language |  | Motor function |  |
| --- | --- | --- | --- | --- | --- | --- | --- | --- | --- |
| CpG | chr | position | Gene | $\Delta$ beta | p-value<sup>a</sup> | $\Delta$ beta | p-value<sup>a</sup> | $\Delta$ beta | p-value<sup>a</sup> |
| cg00490349 | 11 | 45740105 | N/A | -0.031 | 1.94E-05 | **-0.036** | **2.41E-08** | -0.050 | 2.73E-07 |
| cg26971411 | 19 | 41016501 | *SPTBN4* | -0.013 | 3.93E-05 | -0.014 | 2.57E-06 | **-0.024** | **3.10E-08** |

| B. DMRs |  |  |  | Cognition |  | Language |  | Motor function |  |
| --- | --- | --- | --- | --- | --- | --- | --- | --- | --- |
| CpGs | chr | position (start - end) | Genes<sup>b</sup> | Max $\Delta$ beta<sup>c</sup> | p-value<sup>d</sup> | Max $\Delta$ beta<sup>c</sup> | p-value<sup>d</sup> | Max $\Delta$ beta<sup>c</sup> | p-value<sup>d</sup> |
| 9.06E-05 | 1 | 147826191 -<br>147826387 | N/A | N/A | N/A | **-0.099** | **9.06E-05** | N/A | N/A |

$\Delta$ beta: This coefficient represents the mean difference of DNAm beta values between children with and without severe neurodevelopmental delay. Negative coefficients refer to smaller mean DNAm beta values in children with severe neurodevelopmental delay and positive coefficients refer to larger mean DNAm beta values in children with severe neurodevelopmental.
<sup>a</sup> Bonferroni-threshold ( $0.05/403933=1.24e-07$ ) was used to correct for multiple testing in the single CpG analyses. Significant p-values in **bold**;
<sup>b</sup> Annotated genes in the region <sup>c</sup> Maximum $\Delta$ beta for the region; <sup>d</sup> Minimum FDR p-value for the region. Significant p-values in **bold**.

**Figure 1.**
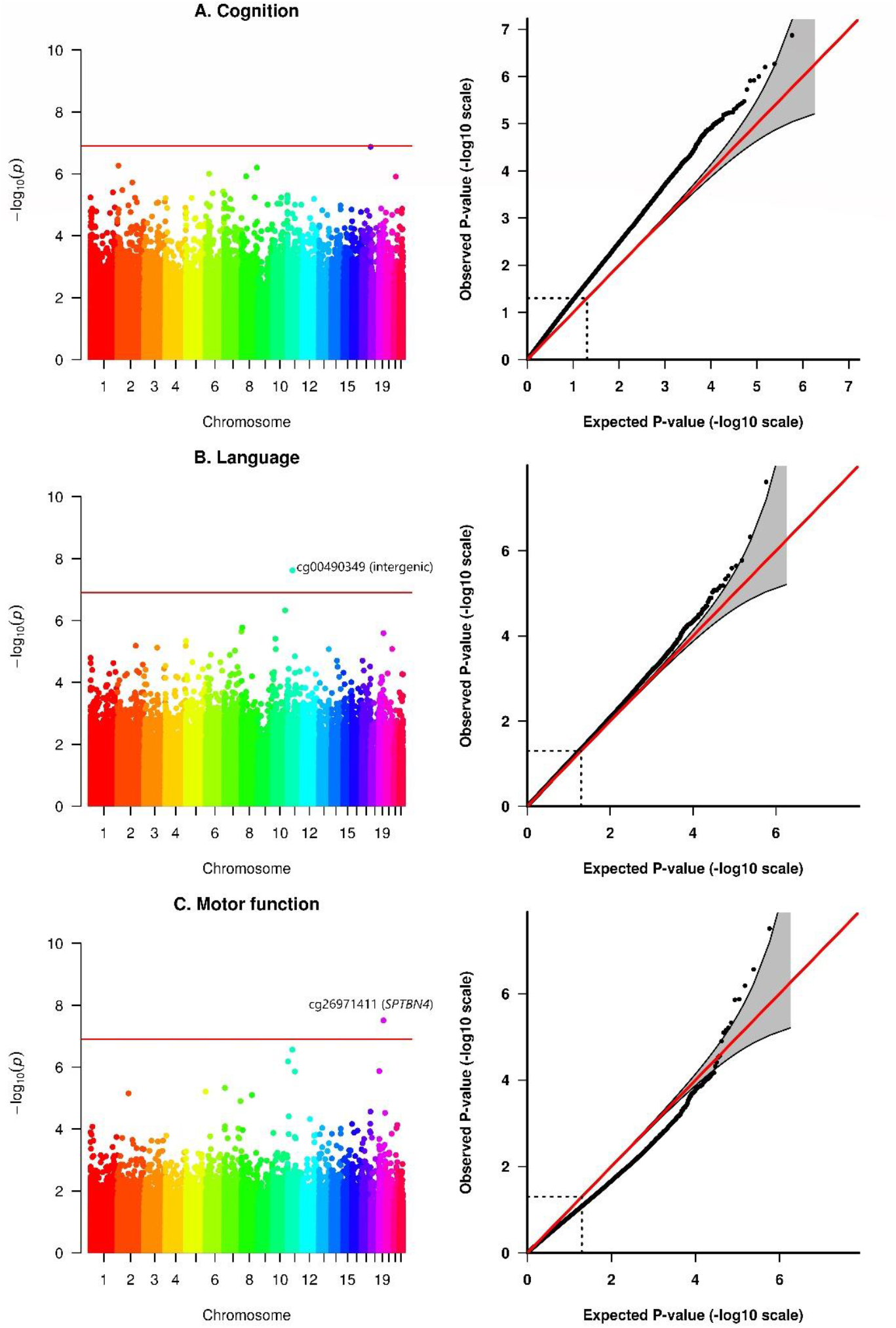
Manhattan and QQ-Plots from the EWAS on severe neurodevelopmental delay in different cognitive domains (A. Cognition, B. Language, C. Motor function). Adjusted for sex, preterm birth, maternal smoking, household income, the first five genetic PCs to control for population stratification and the first three PCs from cell type proportions (after centered log-ratio transformation). Bonferroni-threshold: 0.05/403933 = 1.24 × 10^−07^

In addition to the two significant single CpG sites, we identified one significant DMR from our EWAS results on severe delay in language development that is located in an intergenic region on chromosome 1 (Table 2, maximum **Δ** beta for the region = −0.099, minimum FDR p-value for the region = 9.06 × 10^−05^). Fine mapping of the DMR revealed that it covers a CpG island located in a regulatory region of the genome (promoter and enhancer), which suggests that the methylation state may have an impact on gene expression (Figure S4).

No significantly enriched pathway was found among the most significant CpG sites (p-values < 0.001) from the EWAS of severe neurodevelopmental delay (Tables S3-S5).

### Changes in Neonatal Subcortical Brain Volumes Associated with Neurodevelopmental delay

Increased caudate volumes were associated with severe neurodevelopmental delay at two years of age (Table 3, Figure 2), particularly for language development (**Δ** caudate volume = 165.30, p-value = 0.0443) and delay in motor function (**Δ** caudate volume = 365.36, p-value = 0.0082). The association between caudate volumes and delay in motor function was also significant for mild delay in motor function (**Δ** caudate volume = 156.62, p-value = 0.0148) and when using the continuous Bayley score (p-value = 0.0150) (Figure S5, Table S6). Furthermore, the associations were robust towards additional adjustment for preterm birth, maternal smoking, household income and the first five genetic PCs to control for population stratification (**Δ** caudate volume = 349.97, p-value = 0.0144, Table S7). Associations with language development were only significant for severe delay (Table S6) and not significant after including additional covariates (Table S8). There were no associations with other subcortical brain volumes or with total grey or white matter (Table 3).

**Table 3.**
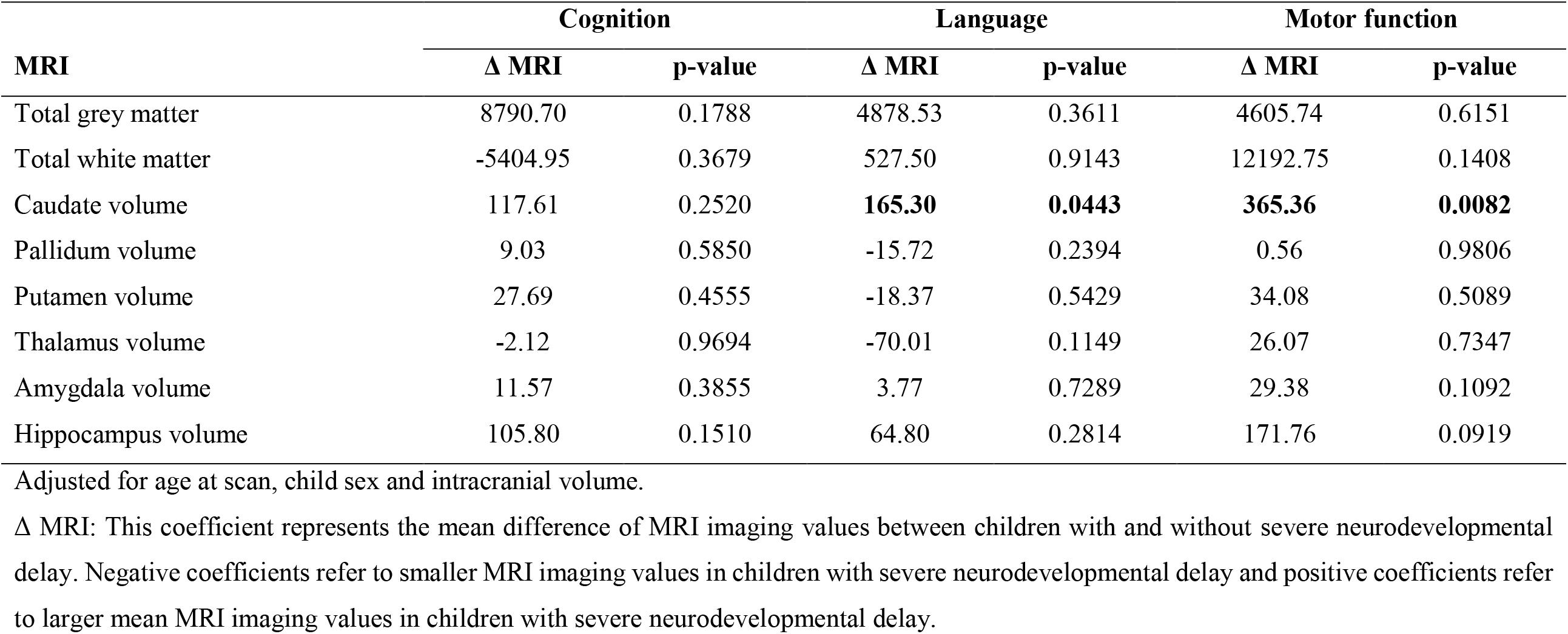
Association between MRI imaging data from neonates and severe neurodevelopmental delay at two years of age.

| MRI | Cognition |  | Language |  | Motor function |  |
| --- | --- | --- | --- | --- | --- | --- |
| | $\Delta$ MRI | p-value | $\Delta$ MRI | p-value | $\Delta$ MRI | p-value |
| Total grey matter | 8790.70 | 0.1788 | 4878.53 | 0.3611 | 4605.74 | 0.6151 |
| Total white matter | -5404.95 | 0.3679 | 527.50 | 0.9143 | 12192.75 | 0.1408 |
| Caudate volume | 117.61 | 0.2520 | **165.30** | **0.0443** | **365.36** | **0.0082** |
| Pallidum volume | 9.03 | 0.5850 | -15.72 | 0.2394 | 0.56 | 0.9806 |
| Putamen volume | 27.69 | 0.4555 | -18.37 | 0.5429 | 34.08 | 0.5089 |
| Thalamus volume | -2.12 | 0.9694 | -70.01 | 0.1149 | 26.07 | 0.7347 |
| Amygdala volume | 11.57 | 0.3855 | 3.77 | 0.7289 | 29.38 | 0.1092 |
| Hippocampus volume | 105.80 | 0.1510 | 64.80 | 0.2814 | 171.76 | 0.0919 |

$\Delta$ MRI: This coefficient represents the mean difference of MRI imaging values between children with and without severe neurodevelopmental delay. Negative coefficients refer to smaller MRI imaging values in children with severe neurodevelopmental delay and positive coefficients refer to larger mean MRI imaging values in children with severe neurodevelopmental delay.

**Figure 2.**
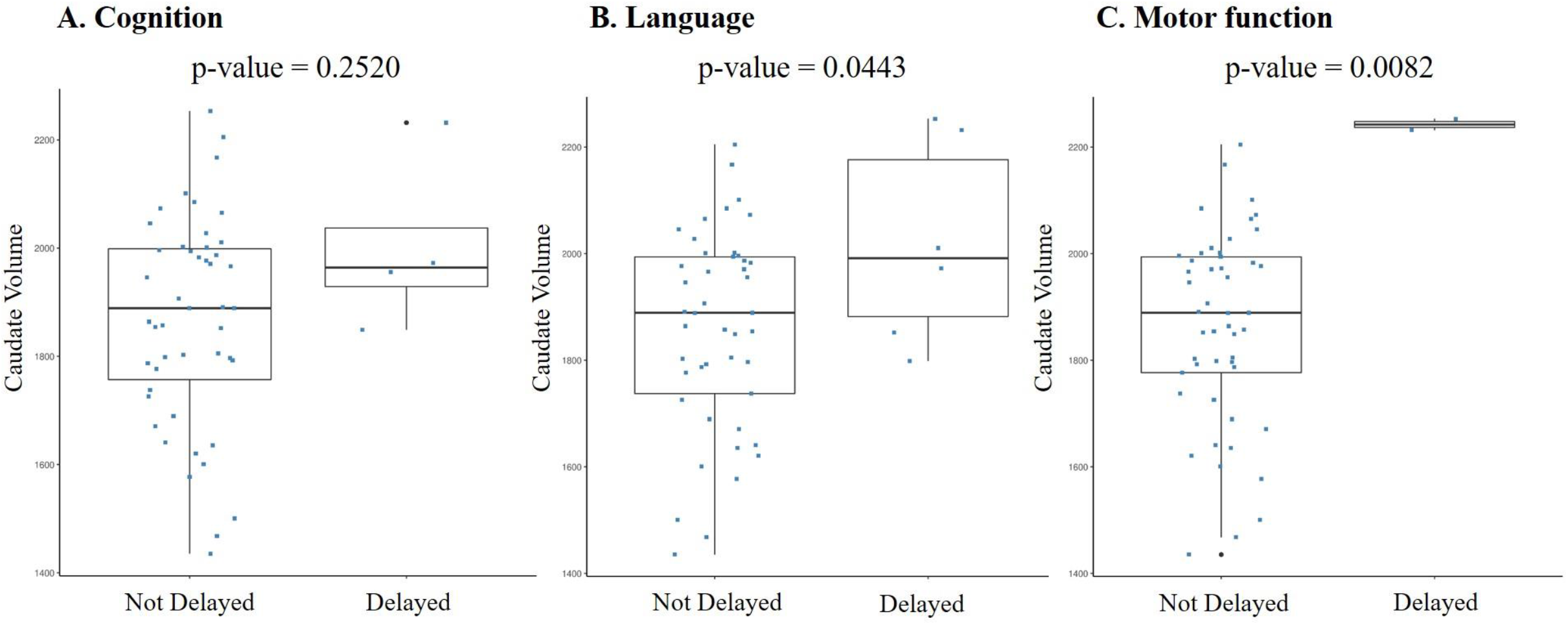
Association between caudate volume in neonates and severe neurodevelopmental delay. Adjusted for age at scan, child sex and intracranial volume.

However, since we did not find associations between methylation in the significant CpG sites from the EWAS of neurodevelopmental delay (cg26971411 and cg00490349) and changes in brain volumes (Table S9), our data did not support a mediating effect of altered brain volumes on the association between methylation and neurodevelopmental delay.

## Discussion

In this study of infants from a peri-urban region in a low-resourced community in South Africa, we showed that differential methylation levels from cord blood were associated with severe neurodevelopmental delay at two years of age. While these associations were not mediated by neonatal brain volume, neonatal caudate volumes were independently associated with severe neurodevelopmental delay, particularly in language and motor domains. These results suggest that methylation levels and brain imaging data from newborns can potentially be useful as unique neurobiological signals for neurodevelopmental delay and highlight the need to understand the biological pathways of how prenatal environmental and psychological risk factors, and genetics affect neurodevelopment. Our findings could have implications for early detection of neurodevelopmental delay and timely implementation of interventions^4^ which are essential for optimal cognitive functioning across the life course^42^.

### Differential Methylation as an Early Sign of Neurodevelopmental Delay

Differential methylation in three loci (*SPTBN4*, an intergenic region on chromosome 11 and a differentially methylated region on chromosome 1) was significantly associated with severe neurodevelopmental delay at two years of age in this cohort. Differential methylation in the *SPTBN4* locus (cg26971411) showed the strongest association with severe delay in motor function. *SPTBN4* (Spectrin Beta, Non-Erythrocytic 4) is a protein coding gene, which is, according to the Human Protein Atlas, primarily expressed in brain tissue and particularly in the cerebellum, the brain region that functions as a co-processor of movement in concert with the cortex and basal ganglia^43^. In line, mutations and loss-of-function variants in *SPTBN4* were reported in association with arthrogryposis, a neuromuscular condition^44^, and with a severe neurological syndrome that includes congenital hypotonia, intellectual disability, and motor axonal and auditory neuropathy^45^. To the best of our knowledge, only one study has identified differential DNAm in *SPTBN4* in association with neurocognitive outcomes (Alzheimer’s disease) and this study was conducted in mice^46^. Therefore, our study extends the current literature by highlighting that not only genetic variants or methylation levels from brain tissue, but also methylation levels from cord blood are linked to cognitive outcomes by providing to our knowledge the first report of an association between neonatal differential methylation in *SPTBN4* in cord blood and early neurodevelopmental outcomes.

The second locus for which we found an association with severe neurodevelopmental delay was a DMR in an intergenic region of chromosome 1. Fine mapping of the DMR revealed that it covers a CpG island located in a regulatory region of the genome (promoter and enhancer), which suggests that the methylation state may have an impact on gene expression. However, more research is needed to determine the genes that are influenced by this region.

### Changes in Neonatal Caudate Volumes Associated with Severe Neurodevelopmental Delay

While we did not find that altered neonatal brain volumes mediate the association between DNAm and neurodevelopmental delay, we demonstrated that larger neonatal caudate volumes were associated with neurodevelopmental delay at two years of age, particularly in motor function. The caudate nucleus is one of the structures that make up the corpus striatum, which is a component of the basal ganglia. The caudate nucleus plays a prominent role in motor processes, and caudate nucleus dysfunction has been found in Parkinson’s disease, Huntington’s chorea, dyskinesias, obsessive–compulsive disorder and other movement and cognitive disorders^47^. Similarly, studies of children have found that the caudate is involved in executive function processes and is associated with autism spectrum disorders^48^. The human brain undergoes rapid change in the first year of life, where the growth rates of subcortical grey-matter structures are similar to cortical grey-matter growth rates, where the amygdala, thalamus, caudate, putamen, and pallidum grow by about 105% in the first year and roughly 15% in the second^49^. Our study finding that larger neonatal caudate volumes were associated with severe neurodevelopmental delay suggest that risk factors for neurodevelopmental delay may impact structural brain development *in utero*, resulting in deviations in maturation that may already be detected soon after birth. This finding suggests the potential use of neonatal brain imaging data as a neurobiological signal for neurodevelopmental delay in addition to DNAm. Here, we assume that any extreme deviation from the normal trajectory may result in adverse cognitive outcomes.

### Strengths & Limitations

Strengths of this study include the very well characterized group of infants as part of a population-based prospective study design with (epi)genetic data from newborns, early MRI scanning in the neonatal period and detailed assessment of neurodevelopment at two years of age. Furthermore, our study population is of African and mixed ancestry, populations which have been understudied in current genetic and epigenetic studies. Beside genome-wide DNAm levels from newborns, our study also included genome-wide genotype data, which we used to correct for population stratification. For this study, 2 to 4-week-old infants were chosen for imaging as the last weeks of gestation and the early postnatal period are a time of marked cerebral maturation. Imaging during the early postnatal period may more accurately reflect the effects of prenatal environmental, psychological risk factors and genetics on brain structure, given the limited exposure to postnatal factors.

Our study has a number of limitations. First, our findings were based on a relatively small sample size, particularly the analyses of neonatal brain imaging data, which may have impacted our results and limited the statistical power to detect mediation effects. Another limitation of our analyses was the small number with severe neurodevelopmental delay in our sample. However, to reduce the risk of false positive findings due to the imbalanced study design, we validated our findings by using different modelling approaches in our EWAS analyses (limma, linear regression, permutation tests, DMR analysis) and different variables to measure neurodevelopmental delay in all our analyses (severe neurodevelopmental delay, mild neurodevelopmental delay, continuous Bayley scores). Although we adjusted our analyses for predefined important covariates, residual confounding cannot be ruled out. Another possible limitation is that we used estimated cell counts in our analyses because measured cell types or single-cell methylation data were not available. However, such estimated cell type adjustments have been shown to be appropriate in epidemiological settings^50^. Methylation signatures are tissue and cell-type specific, and therefore, selection of relevant tissues and cell-type is of crucial importance for epigenetic analyses. While brain tissue would be the most relevant tissue in association with neurodevelopment, we analyzed methylation levels in cord blood, which has the advantage that it is easily assessible from living samples and could therefore be linked to future cognitive outcomes. Furthermore, we identified differentially methylated loci in cord blood which were mapped to a known candidate gene for several related cognitive outcomes^44–46^ and a regulatory region of the genome (promoter and enhancer), strengthening the validity of our findings.

## Conclusions

We have presented evidence that differential neonatal methylation in *SPTBN4*, a regulatory region on chromosome 1, and an intergenic region on chromosome 11, as well as increased neonatal caudate volumes, were independently associated with severe neurodevelopmental delay at two years of age. These findings suggest that neurobiological signals for severe developmental delay may be detectable in very early life with implications for identification and intervention design.

## Data Availability

Data are available upon request from the Steering Committee (Prof. Heather Zar)

## Acknowledgments

The authors thank the study and clinical staff at Paarl Hospital, Mbekweni and TC Newman clinics, as well as the CEO of Paarl Hospital, and the Western Cape Health Department for their support of the study. The authors thank the families and children who participated in this study.

